# Comparison of Structural Diagnosis and Management (SDM) Approach versus Myofascial Release (MFR) for Plantar Heel Pain in People with Diabetes Mellitus: A Study Protocol for a Multicentre Randomised Clinical Trial

**DOI:** 10.1101/2025.09.07.25335270

**Authors:** Paroshmoni Biswas Mim, Suvro Nill Sarkar, Kazi Md Azman Hossain, Md. Feroz Kabir, Md. Zahid Hossain, Sharmila Jahan, Ehsanur Rahman, Abid Hasan Khan, K M Amran Hossain

## Abstract

**Introduction:** Plantar heel pain (PHP) is a common musculoskeletal condition among individuals with diabetes mellitus (DM), often linked to plantar fasciitis, heel spurs, and myofascial dysfunction. It leads to chronic pain, limited mobility, and reduced quality of life. Although manual therapy is effective in non-diabetic populations, there is limited evidence supporting its use in diabetic patients. This multicentre randomised clinical trial aims to compare the effectiveness of Structural Diagnosis and Management (SDM) versus Myofascial Release (MFR) in managing PHP in people with diabetes mellitus, with a focus on reducing pain and improving function and disability outcomes to address a critical clinical and research gap.

**Methods and analysis:** This 8-week, parallel-group, assessor- and participant-blinded, two-arm randomised clinical trial will include 90 diabetic participants aged 30–65 years with PHP, recruited from three diabetic hospitals in Bangladesh. Participants will be randomly assigned (1:1) to receive either the SDM or MFR therapeutic approach, with 24 sessions over 8 weeks (each lasting 45–60 minutes). The primary outcome will be pain intensity, measured using a 10 cm Visual Analogue Scale (VAS). Secondary outcomes will include ankle range of motion, muscle strength, and the Foot Function Index (FFI). Assessments will be conducted at baseline, posttest (8 weeks), and follow-up (20 weeks) to evaluate both immediate and long-term effects.

**Ethics and dissemination:** The study received ethical approval from the Institutional Review Board of the Department of Physiotherapy and Rehabilitation, Jashore University of Science and Technology (Approval ID: PTRJUST/IRB/2025/03/192411). All activities and exercise programs will be conducted following the 2020 Helsinki Declaration. Findings will be shared through professional conferences and in an international, Scopus-indexed, peer-reviewed journal to inform clinical guidelines for PHP management in individuals with diabetes mellitus.

**Trial registration number:** The Trial was Registered Prospectively. Trial number was CTRI/2024/11/076311 [Registered on: 05/11/2024]

**What is already known on this topic?:** Plantar heel pain is common among people with diabetes mellitus, causing pain, limited mobility, muscle weakness, and reduced function. Although various treatments are available, many lack long-term effectiveness and value for money. SDM and MFR are specific, evidence-based physiotherapy methods; however, there is limited comparative evidence on their efficacy in this population.

**What this study adds?:** This study presents a direct comparison between the SDM and MFR approaches in managing plantar heel pain in individuals with diabetes mellitus. It will assess key clinical outcomes—pain intensity, ankle range of motion, muscle strength, disability, and activity limitation—over an 8-week intervention period, followed by a 12-week follow-up. The study aims to establish evidence for a more effective and sustainable physiotherapy intervention, thereby improving treatment protocols in this population.

**How this study might affect research, practice or policies?:** By determining the most effective intervention between SDM and MFR, this study will promote evidence-based physiotherapy practices for managing plantar heel pain in diabetic patients. The findings may inform clinical decision-making, contribute to the development of cost-effective, non-pharmacological treatment guidelines, and influence future research directions and rehabilitation policies in diabetic foot care.

## INTRODUCTION

Plantar heel pain (PHP) is the most common foot condition affecting the plantar region of the foot and is associated with surrounding structures, including the calf muscle, associated nerves, and plantar fascia, in individuals with a sedentary lifestyle or those who are active and maintain a prolonged standing or walking posture. Clinically, PHP can be associated with plantar fasciitis, typically characterised by discomfort, intermittent pain, and tenderness in the area of fascia located underneath the heel, extending towards the medial longitudinal arch (MLA) of the foot. There are two common clinical conditions associated with PHP, where plantar fasciitis is the most prevalent cause of plantar heel pain.^1^ The thick band of connective tissue called plantar fascia, which consists of central, medial, and lateral bands. Where the central band arises from the medial tubercle of calcaneal bone toward toes, which provides support for the foot arch, that tissue becomes irritated and inflamed due to tensile overload or force. Calcaneal spur is another cause. It is a fibromatosis that progressively infiltrates the plantar aponeurosis due to mechanical stress on the foot, gradually and continually increasing in the insertion point.^2^ A heel spur is radiologically evident, and pain eventually extends from the heel to the lower extremities, worsening with walking or during the first minutes of rest. Painful ambulation leads to functional impairments. Moreover, this recurrent chronic condition hinders quality of life for an individual and presents with noteworthy disability.^3^

The study of PHP incidence and prevalence is limited. It is estimated that 10% of individuals with lower limb pain can experience this condition in their lifespan.^4, 5^ An epidemiological study in the USA has demonstrated that nearly one million individuals visit physicians annually for PHP.^6^ Risk factors of PHP include obesity, decrease the arches, reduce flexibility, pes planus, improper footwear or hard surfaces of footwear, leading sedentary lifestyle, and prolong working posture in walking or standing barefoot on the hard surfaces or floors.^7^ Strong association has been noted with PHP and metabolic disease such as diabetes mellitus, hyperuricemia, and hyperglycaemia. The prevalence of PHP is higher among people with diabetes mellitus (DM) and diabetic foot (DF).^8, 9^

Systematic review between (1994 and 2013) reports, 4.5% to 35.0% of Bangladeshi people have DM. The number might rise to 13.7 million by 2045.^10^ Approximately one million individuals with DM visit physicians for PHP.^11^ Globally, 0.85% of diabetic people have PHP; among them, 1.31% have type-2 and 0.92% have type-1 DM.^12^ The clinical presentation and biomechanical factors of PHP are similar in both non-diabetic and diabetic patients. However, in DM cases with PHP, elderly women with a high BMI are the most vulnerable group. In DM cases, degeneration of the plantar fascia and repetitive micro-tears cause relapsing inflammation and delayed healing, creating a vicious cycle of injury and pain. Shortening of the calf muscle leads to a series of biomechanical abnormalities, such as a decrease in the range of motion (ROM) of dorsiflexion and an increase in stress to the perifascial structures. Diagnosis of PHP can be made based on plantar fascia, heel spur, or myofascial abnormalities of the foot, according to the International Statistical Classification of Diseases and Related Health Problems (ICD) category. Clinical features are categorised according to specific diagnosis. Some clinical examination, such as the windlass test, longitudinal arch angle, tarsal tunnel syndrome test, and active and passive dorsiflexion ROM in the talocrural joint, supports the diagnosis.^13, 14^

Intervention guidelines for PHP in non-diabetic and diabetic patients are reported similarly in the previous clinical practice guidelines. Medications include anti-inflammatory agents, such as NSAIDs. Many studies reported the significance of steroid injections in PHP in non-diabetic patients and some in diabetic patients to reduce inflammation, improve pain, and functional conditions. However, steroids have a long-term consequence of tendon and osseous degeneration. In physiotherapy practice, various interventions have been reported, including electrical modalities such as iontophoresis with dexamethasone 0.4% or acetic acid 5%, which give pain relief and improve function within 2 to 4 weeks. Manual therapy techniques such as nerve mobilization (effective within 1 to 3 months), soft tissue mobilization, passive neural mobilization, gliding of the talocrural joint posteriorly, the subtalar joint laterally, and the first tarsometatarsal joint anterior-posteriorly, and distraction manipulation of the subtalar joint are also commonly used to reduce pain and restore mobility for people with PHP. Other conservative approaches include taping, orthotic devices, and night splints. Calcaneal taping may provide pain relief temporarily within 7 to 10 days. Orthotic devices, such as specially designed foot orthoses, reduce pain and improve function over a short-term period of approximately three months. Night splints, available in sock-type, anterior, or posterior designs, are recommended for individuals with plantar heel pain lasting six months or more. They are typically worn for 1–3 months.^15^ Studies on PHP in non-diabetic patients have revealed that myofascial release to the calf muscles and plantar fascia is a practical manual therapy technique.^16^ Recent studies indicate that the Structural Diagnosis and Management (SDM) approach—targeting calf muscles, plantar fascia, lower limb muscles, and myoneural structures—offers short- to intermediate-term pain relief and improved functional independence in people with PHP. However, PHP is higher in DM cases and has a poor prognosis, and there is a significant research gap addressing effective manual therapy approaches for PHP in people with DM.^17–21^ Therefore, the primary goal of this study is to compare the effectiveness of two manual therapy approaches, Structural Diagnosis and Management (SDM) and Myofascial Release (MFR), in treating individuals with PHP who have DM. The objectives are (1) to determine baseline compatibility of both groups in socio-demographic and clinical variables; (2) to measure between-group and within-group post-treatment outcomes in pain, ankle ROM, muscle strength, activity limitation, and disability; and (3) to compare the follow-up outcomes of both groups. After 8 weeks of interventions, the SDM or MFR group is expected to show better improvement in PHP for people with DM.

## METHODS

### Study design

A multicentre, parallel, assessor- and participant-blinded randomised clinical trial will be conducted with 8 weeks of interventions at Nurul Islam Diabetic Centre, Ibn Sina Diabetes Centre, and City Hospital & Diabetic Care, Jashore, in Bangladesh, from September 2025 to March 2026. The eligible participants will be randomly allocated following a 1:1 ratio to either the SDM or MFR group. This trial has been designed in accordance with the Standard Protocol Items: Recommendations for Interventional Trials (SPIRIT) guidelines, which provide a comprehensive framework for the development of RCT protocols, as detailed in Table 1. Additionally, the Consolidated Standards of Reporting Trials (CONSORT) statement has been followed to enhance the quality and completeness of RCT reporting, as detailed in Figure 1 (CONSORT flow diagram) which is attached in the supplementray file.

**Table 1:**
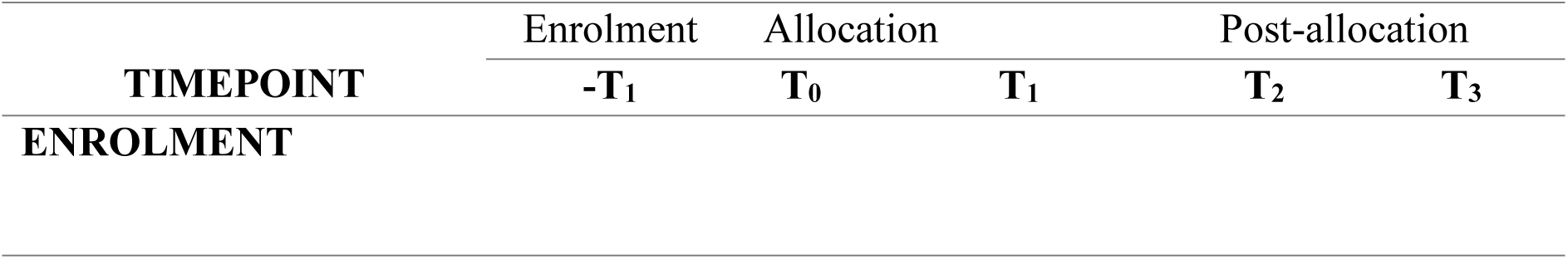
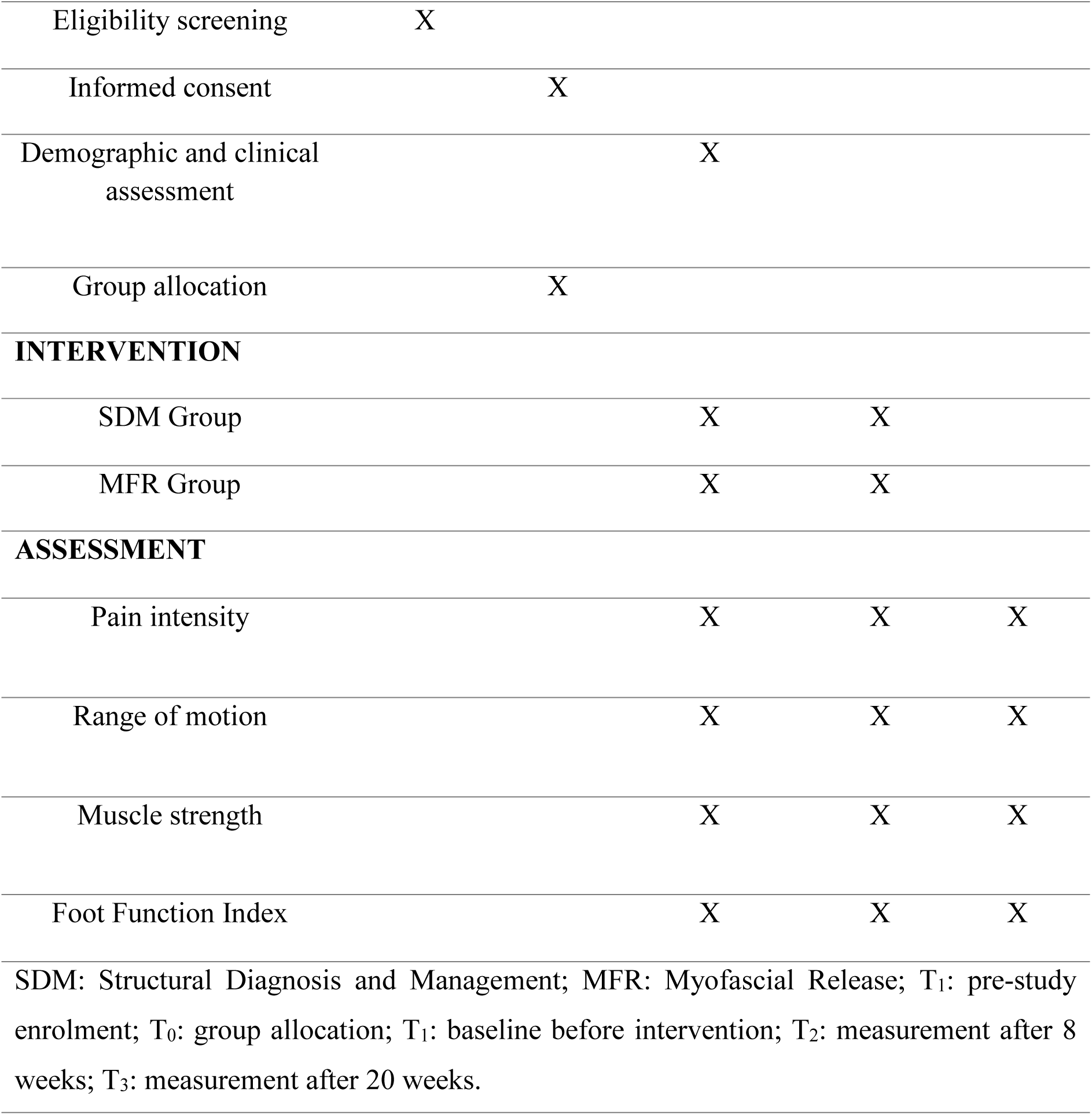
SPIRIT table.

**Table 2:**
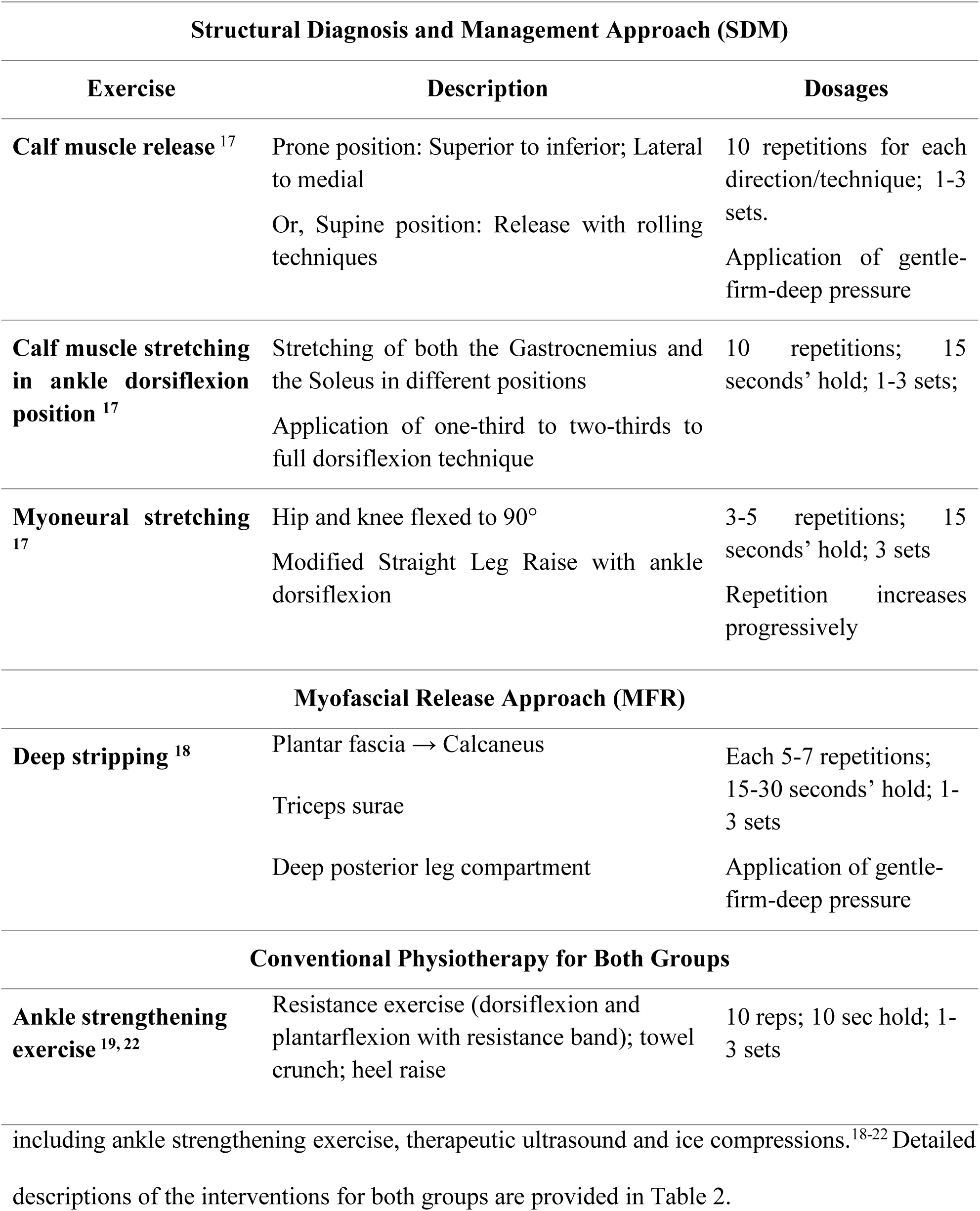
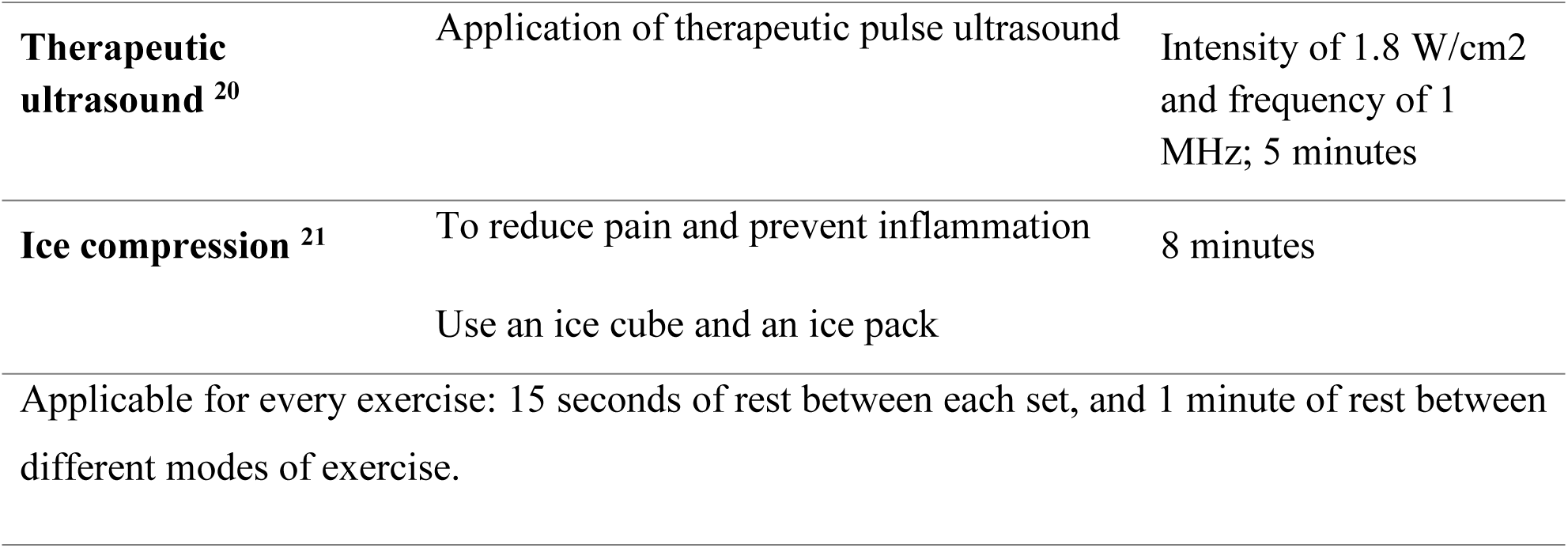
Intervention Details.

**Figure 1:**
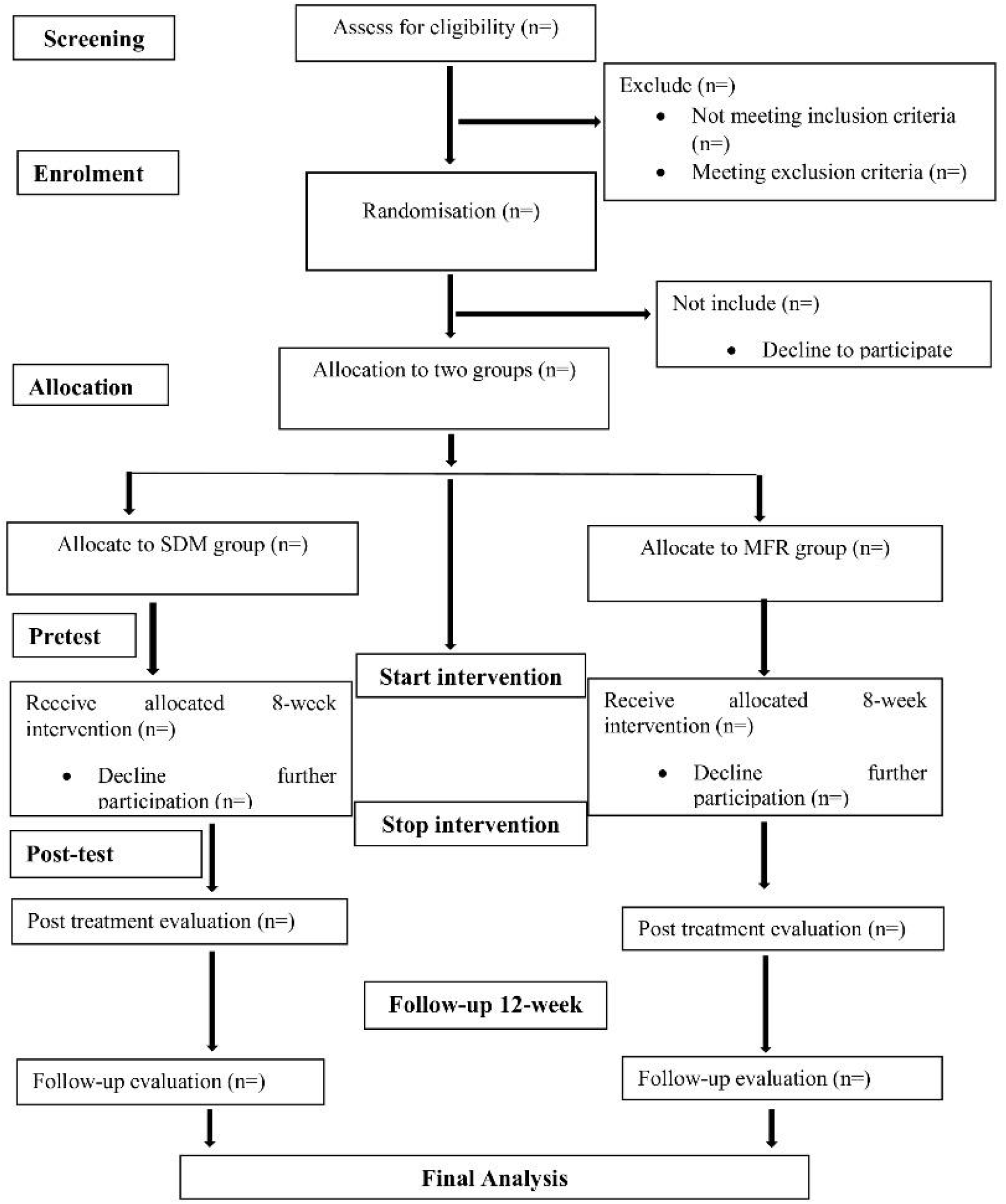
CONSORT flow diagram.

### Sample size calculation

The required sample size was calculated using G*Power software version 3.1.9.7 (University of Kiel, Kiel, Germany). Based on an effect size of 0.50, with a significance level (α) of 0.05 and statistical power of 0.80, a total of 72 participants were required for the study. To assume a potential 25% dropout rate, the final adjusted sample size was increased to 90 participants, with 45 allocated to each group.^3^

### Participant recruitment and screening

Between September 2025 to March 2026, DM with PHP will be recruited from three hospitals in Jashore, Bangladesh. Participants will be screened for eligibility by two trained research assistants, and those who meet the criteria will provide written informed consent. Following CONSORT guidelines, participants will be randomly assigned to their allocated groups using a concealed allocation method. Four blinded assessors will conduct baseline assessments prior to the intervention’s commencement. The intervention will follow a standardised, concealed protocol, with participants blinded to group allocation to minimise bias.

### Eligibility criteria

Participants will be screened for eligibility by trained research assistants. Inclusion criteria: (1) age between 30-65 years for both male and female;^17^ (2) participants diagnosed with unilateral or bilateral heel pain or plantar fasciitis (according to ICD-10 criteria, ICD diagnosis codes M72.2); (3) localized pain in the plantar region of the foot when placed their barefoot on the floor in the morning; (4) ankle dorsiflexion is more limited than plantar-flexion; and (5) suffering with plantar heel pain from more than 4 weeks.^17^ Exclusion criteria: (1) history of lower extremity fracture, rheumatoid arthritis, osteoarthritis, carcinoma, surgery of the foot, and metal implant around ankle;^15^ (2) active foot ulceration, infection, severe peripheral vascular disease, gangrene of the foot, cyanotic symptom, and severe neurological condition;^11^ (3) contraindicated to manual therapy or exercise intervention;^17^ (4) participants taken corticosteroids injection within 6 months.^16^

### Randomisation and blinding

Eligible participants who will give written informed consent and will be randomly allocated to their respective groups, either the SDM or the MFR group. The randomisation sequencing process will be conducted using the “Rand” function in Microsoft Excel 2013 software. A sequential list of participants’ IDs, sorted according to the randomly generated numbers. This procedure helps to prevent bias. Participants will be allocated to a parallel intervention group in a 1:1 ratio. This will be a double-blind, randomised clinical trial, where both participants and assessors will maintain blinding to prevent bias and data contamination, while physiotherapists delivering the treatments will be fully informed about the interventions.

### Interventions

Both groups will undergo an 8-week intervention program, consisting of three sessions per week, with each session lasting 45 to 60 minutes, following the designed treatment protocol.

The SDM group will receive manual therapy based on the Structural Diagnosis and Management (SDM) approach for PHP. This technique targets the calf muscles using graded interventions classified by pressure velocity and biomechanical positioning, combined with stretching of myoneural structures.^17^ The MFR group will receive Myofascial Release therapy, involving deep stripping of the plantar fascia and targeted surface points of the triceps surae, following a standard protocol.^18^ Both groups will receive designed conventional physiotherapy treatment

The duration and frequency of sessions may be adjusted individually based on each participant’s condition. To ensure accurate tracking, a monitoring sheet will be maintained for each participant attending every session.

### Outcome measurements

Baseline socio-demographic and clinical characteristics will be collected immediately after obtaining informed consent from participants and before the initiation of the intervention. All primary and secondary outcomes will be analysed at three time points: baseline (week 0), posttest (week 8), and follow-up (week 20), by four trained assessors who will be blinded to group allocation.

### Socio-demographic and clinical characteristics

The researcher will develop a structured questionnaire to collect socio-demographic and clinical characteristics from participants. This will include personal details such as age, gender, and occupation; demographic information; and condition-related variables such as BMI, type and duration of DM, radiological confirmation of diagnosis, duration of PHP, affected foot, type of footwear, and duration of footwear use.

### Primary outcome

#### Pain intensity

Pain intensity will be measured using a 10 cm Visual Analogue Scale (VAS), a widely accepted tool for evaluating pain. The scale consists of a 10 cm horizontal line anchored at 0 ("no pain") and 10 ("severe pain"). The VAS demonstrates excellent test-retest reliability (ICC = 0.99; 95% CI: 0.989–0.992) and strong validity.^23^

#### Secondary outcomes Range of Motion (ROM)

Participants will actively perform dorsiflexion and plantarflexion using a digital dynamometer (ActiveForce-2), with measurements recorded in degrees via the device’s app. A study reported excellent reliability for the portable dynamometer, with ICC values of 0.972 for dorsiflexion and plantarflexion. Additionally, strong reliability was found between right and left ankle measurements: plantarflexion ICC = 0.89 and dorsiflexion ICC = 0.94.^24, 25^

#### Muscle strength

A portable digital hand dynamometer (ActiveForce-2) and its companion app will be used to assess isometric strength of the dorsiflexors and plantarflexors of the affected foot, with measurements recorded in kilogram-force (kgf). Previous research has demonstrated that portable dynamometers exhibit high inter-rater and test-retest reliability, with strong correlations for dorsiflexion and plantarflexion strength (r = 0.827, r = 0.973).^25^

#### Disability and activity limitation

To assess overall pain, disability, and activity limitation, the Foot Function Index (FFI) will be used. The FFI is a self-administered questionnaire comprising 23 items, divided into three subscales. According to previous studies, the FFI has shown test-retest reliability ranging from 0.69 to 0.87 and internal consistency between 0.73 and 0.96. It’s strong correlation with clinical measures of foot pathology will support its validity as a standard tool for this study. ^26, 27^

#### Study procedure

After screening by trained research assistants, eligible participants will be thoroughly informed about the study’s objectives and procedures, and then obtain their written informed consent. Once screened, participants will be randomly assigned to their allocated groups using a predetermined randomisation sequence to ensure unbiased allocation. Four blinded assessors will then collect baseline data at the study centre using an ethically approved questionnaire designed specifically for this research, which includes socio-demographic information and clinically relevant measures to confirm baseline compatibility with no statistically significant differences between groups. Following the baseline assessment, participants will undergo their assigned intervention for 8 weeks. After this period, blinded assessors will perform posttest (week 8) and follow-up (week 20) evaluations by measuring primary and secondary outcome variables. This entire study procedure will strictly follow the Standard Protocol Items: Recommendations for Interventional Trials (SPIRIT) 2013 guidelines (table 1), ensuring methodological rigour and ethical compliance throughout the study.

#### Safety measures and adverse reaction management

Although no adverse events are anticipated from the treatment, the monitoring team remains vigilant and will promptly notify the appropriate healthcare professionals of any unexpected occurrences that may arise during or after the intervention. All treatment procedures will be thoroughly documented in daily SOAP notes to ensure clear records and to prevent any future misunderstandings or complaints from participants. Before initiating the intervention, the physiotherapist will carefully screen for any contraindications. Should participants experience any discomfort, pain, or skin irritation after the treatment, they should immediately report these symptoms to both the physician and physiotherapist. Identified any harmful effects, which will be transparently reported by the researcher in the final publication. Additionally, an adverse event reporting checklist will be provided throughout the intervention period to facilitate systematic monitoring.

### Data Management

After each assessment, data will be verified daily by the assessors to ensure accuracy. The final study datasets will be accessible only to the study manager, principal investigators, and data auditors. Upon completion of the study, all authors will have equal access to anonymised data. The principal investigator will securely store all hard and electronic copies of the collected data, ensuring that no access is granted to identifiable participant information. Manual data entries will undergo double-checking to minimise errors. Online data will be carefully stored on a password-protected server managed by the postgraduate program in Physiotherapy and Rehabilitation. Each participant will be assigned a unique identification number, and data will be encoded securely using this ID. The ID number will be encrypted separately from any identifying information to maintain participant confidentiality throughout the study.

### Data monitoring

Two independent individuals will be appointed to monitor and audit group enrollment, the intervention protocol, and any adverse effects experienced by participants. They will not be directly included in this study. These monitors will have access to review data and conduct interim analyses. Any changes to the research or treatment protocol will be promptly communicated by the principal investigator to the Ethical Review Board.

### Data analysis

Data analysis will be performed by a statistician using SPSS version 22 with encrypted data to ensure confidentiality. The parametricity of the data will be assessed using the Kolmogorov-Smirnov and Shapiro-Wilk tests, as well as measures of skewness and kurtosis, and visualisation of the bell curve. Descriptive statistics, including mean, standard deviation, median, interquartile range, frequency, and percentage, will summarise continuous and categorical variables. For comparisons across three time points, repeated measures ANOVA will be used for normally distributed data, while the Friedman test will be applied as the non-parametric alternative. Between-group and within-group differences will be analysed using independent and paired t-tests for parametric data, or Mann-Whitney U and Wilcoxon signed-rank tests for non-parametric data. Significance will be set at *p* < 0.05. Baseline, posttest and follow-up change scores will also be evaluated, with intention-to-treat analysis applied to address missing data.

### Dissemination

The results of this study will be submitted for publication to peer-reviewed journals. Initially, a seminar will be organised to present the findings to physiotherapists, researchers, and healthcare professionals. Following this, a training and knowledge-sharing session will be conducted specifically for physiotherapists to enhance their understanding of the treatment approach. Disseminating research findings is a crucial part of the research process, facilitating knowledge exchange among researchers, clinicians, healthcare providers, and broader audiences. This process also helps identify gaps for future research. Finally, the study’s results will be presented at national and international conferences worldwide.

## DISCUSSION

The primary aim of this study is to evaluate the effectiveness of the Structural Diagnosis and Management (SDM) Approach versus Myofascial Release (MFR) in treating plantar heel pain in diabetic patients. Previous research indicates that approximately one million individuals are referred annually to a doctor for a definitive diagnosis of plantar fasciitis. A recent cohort study identified the prevalence of plantar fasciitis as 720,000, with an incidence of 0.85% among diabetic patients.^8–12^ Several studies have explored various interventions that could provide interim relief from plantar heel pain, plantar fasciitis, and heel spurs, such as considering and providing anti-inflammatory agents, electrical modalities, and manual therapies like nerve mobilisation, often focusing solely on the central pain area rather than the affected structures.^14, 15^

However, our study considers the potential scope by including the associated structures, which may be a more critical factor in achieving sustainable outcomes. Without addressing these, diabetic patients are increasingly at risk of this foot condition, which often remains untreatable and misunderstood. To date, no research has directly compared and analysed yet the effectiveness of SDM versus MFR in treating plantar heel pain in people with diabetes mellitus. The long-term objective of this study is to play a vital role in the management of this global condition. This study offers several significant benefits. It marks the first attempt in Bangladesh to compare the effectiveness of Structural Diagnosis and Management (SDM) and Myofascial Release (MFR) in managing plantar heel pain among individuals with diabetes mellitus, incorporating a 12-week follow-up period. The study highlights the potential contribution of these intervention strategies not only in reducing pain but also in improving functional outcomes that enhance patients’ quality of life. By addressing a significant gap in existing research, this study may offer valuable insights for clinical practice and future research in diabetic foot management. However, limitations include the focus on a specific area in Bangladesh, which may restrict the broader applicability of its findings. Additionally, the presence of multiple comorbidities and a limited sample size are notable constraints. Future research should involve a larger and more diverse population, with an extended follow-up duration. It is also recommended that subsequent studies adopt more rigorous research designs, suited to resource-limited settings, to enhance the validity and generalisability of the findings.

## Study Status

This randomised clinical trial has not yet recruited any participants.

## Supporting information

SPIRIT checklist

Consent Form in English

CTRI_TRIAL. (prospectively reg.)

EthicsApproval_PTR JUST_2025

## Data Availability

No data are available as no datasets were generated or analysed for this study. After completing the final results of this study, the datasets of the participants and the statistical analysis will be made available to the corresponding author upon reasonable request.

## Acknowledgements

The authors sincerely acknowledge the invaluable support of the authorities, clinicians, and outcome assessors at Nurul Islam Diabetic Centre, Ibn Sina Diabetes Centre, City Hospital & Diabetic Care, Jashore, Bangladesh, as well as the Department of Physiotherapy and Rehabilitation, Jashore University of Science and Technology. We also extend our gratitude in advance to the future participants who consent to take part in this study.

## Contributors

Conceptualisation: PBM, KMAH. Methodology: PBM, KMAH, and KMAH. Software: MFK and MZH. Investigation: SNS, and SJ. Writing original draft: PBM, KMAH, AHK, and KMAH. Writing review and editing: PBM, SNS, KMAH, MFK, MZH, SJ, ER, AHK, and KMAH. Visualisation: ER and KMAH. Supervision: KMAH. Project administration: KMAH, MFK, MZH, SJ, ER, and KMAH. Guarantor: KMAH.

## Patient and public involvement

Patients and/or the public were not involved in the design, conduct, reporting, or dissemination plans of this research.

## Patient consent for publication

Not applicable.

## Ethics approval

This study received ethical approval from the Institutional Review Board, Department of Physiotherapy and Rehabilitation, Jashore University of Science and Technology (Approval ID: PTRJUST/IRB/2025/03/192411). It is also prospectively registered with the Clinical Trial Registry India (CTRI) (Registration ID: CTRI/2024/11/076311) [Registered on: 05/11/2024]. The study will follow the 2020 Declaration of Helsinki. Participant rights will also be protected in accordance with the World Medical Association’s ethical guidelines.

## Provenance and peer review

Not commissioned; internally peer-reviewed.

## Supporting information

**Supplementary file 1. SPIRIT Checklist**

(DOC)

**Supplementary file 2. Ethical Approval Letter**

(PDF)

**Supplementary file 3. Patient Consent Form**

(DOC)

**Supplementary file 4. Funding Letter**

(PDF)

