## Supplementary material for "Comparison of Structural Diagnosis and Management (SDM) Approach versus Myofascial Release (MFR) for Plantar Heel Pain in People with Diabetes Mellitus: A Study Protocol for a Multicentre Randomised Clinical Trial": Consent Form in English

### Patient Consent Form

Assalamu Alaikum / Namaskar,

I am Paroshmoni Biswas Mim, an intern physiotherapist of the Department of Physiotherapy and Rehabilitation, Jashore University of Science and Technology, Jashore 7408. I am conducting a research project under the guidance of Dr. Kazi Md. Amran Hossain, Lecturer of Physiotherapy and Rehabilitation, Jashore University of Science and Technology. My research topic is "Comparison of Structural Diagnosis and Management (SDM) Approach versus Myofascial Release (MFR) for Plantar Heel Pain in People with Diabetes Mellitus: A Multicenter Randomized Clinical Trial." The studies have Ethical approval (ID: PTR-JUST/IRB/2025/03/192411).

This study is an experimental study, and if you are interested in participating, you will be asked a few questions. You can leave or withdraw from the questionnaire at any time during the question period. This data will be kept safe and not provided to anyone without the patient's permission. It can take you 20-25 minutes to complete the entire question paper. Follow the instructions in the questionnaire. If you need any help writing the answer, feel free to take it. We hope that through this research, we can compare the Structural Diagnosis and Management (SDM) Approach and Myofascial Release (MFR) for Treating Plantar Heel Pain in Diabetic Patients.

If you have something to know about this research, you can find out from me on the phone number
(01312-039527). If you want to know further about this study, you can contact my supervisor (mail: ) (phone: 01735661492).

Are you willing to participate in this research study voluntarily? (Put a tick mark): Yes/No

Move forward if you have:

Patient's ID No: ……………………………………..…

Name of Participant: …………………………………………………….

Address: ………………………………………………………………….

Participant's Contact Number: …………………………………………

Participant's Signature: …………………………………………………

Researcher's Signature: …………………………………………………

Date of informed consent process: …………………………………….
