## Supplementary material for "Comparison of Structural Diagnosis and Management (SDM) Approach versus Myofascial Release (MFR) for Plantar Heel Pain in People with Diabetes Mellitus: A Study Protocol for a Multicentre Randomised Clinical Trial": CTRI_TRIAL. (prospectively reg.)

### Clinical Trial Details (PDF Generation Date :- Fri, 20 Dec 2024 03:55:49 GMT)

| | |
|---|---|
| **CTRI Number** | CTRI/2024/11/076311 [Registered on: 05/11/2024] - **Trial Registered Prospectively** |
| **Last Modified On** | 20/12/2024 |
| **Post Graduate Thesis** | No |
| **Type of Trial** | Interventional |
| **Type of Study** | Physiotherapy (Not Including YOGA) |
| **Study Design** | Randomized, Parallel Group Trial |
| **Public Title of Study** | Structural Diagnosis and Management Approach and Myofascial Release for Heel Pain in Diabetic Patients. |
| **Scientific Title of Study** | Comparison of Structural Diagnosis and Management Approach and Myofascial Release for Plantar Heel Pain in Diabetic Patients. |

**Secondary IDs if Any**

| Secondary ID | Identifier |
|---|---|
| NIL | NIL |

**Details of Principal Investigator or overall Trial Coordinator (multi-center study)**

| Details of Principal Investigator | |
|---|---|
| **Name** | Paroshmoni Biswas Mim |
| **Designation** | Undergratuate student |
| **Affiliation** | Jashore University of Science and Technology, |
| **Address** | Bachelor of Physiotherapy Department of Physiotherapy and Rehabilitation Jashore University of Science and Technology Jashore-7408<br><br>7408<br>Other |
| **Phone** | 8801312039527 |
| **Fax** | |
| **Email** | |

**Details Contact Person (Scientific Query)**

| Details Contact Person (Scientific Query) | |
|---|---|
| **Name** | Dr Kazi Md Amran Hossain |
| **Designation** | Lecturer |
| **Affiliation** | Jashore University of Science and Technology |
| **Address** | Lecturer Department of Physiotherapy and Rehabilitation Jashore University of Science and Technology Jashore-7408 Bangladesh.<br><br>7408<br>Other |
| **Phone** | 8801735661492 |
| **Fax** | |
| **Email** | |

**Details Contact Person (Public Query)**

| Details Contact Person (Public Query) | |
|---|---|
| **Name** | Dr Kazi Md Amran Hossain |
| **Designation** | Lecturer |
| **Affiliation** | Jashore University of Science and Technology |
| **Address** | Lecturer Department of Physiotherapy and Rehabilitation Jashore University of Science and Technology Jashore-7408 Bangladesh.<br><br>7408<br>Other |

| | |
|---|---|
| **Phone** | 8801735661492 |
| **Fax** | |
| **Email** | |

**Source of Monetary or Material Support**

| Source of Monetary or Material Support |
|---|
| > MSK Lab - Room No: 308, Department of Physiotherapy and Rehabilitation, Jashore University of Science and Technology, Jashore-7408, Bangladesh |

**Primary Sponsor**

| Primary Sponsor Details | |
|---|---|
| **Name** | Paroshmoni Biswas Mim |
| **Address** | Bachelor of Physiotherapy (BPT), Department of Physiotherapy & Rehabilitation Jashore University of Science & Technology (JUST),Jashore, Bangladesh. |
| **Type of Sponsor** | Other [self-funded] |

**Details of Secondary Sponsor**

| Name | Address |
|---|---|
| NIL | NIL |

**Countries of Recruitment**

| List of Countries |
|---|
| Bangladesh |

**Sites of Study**

| Name of Principal Investigator | Name of Site | Site Address | Phone/Fax/Email |
|---|---|---|---|
| Dr Md Kabir Hossain Clinical Consultant PTR Dept JUST | Dept of Physiotherapy and Rehabilitation, JUST, Jashore-7408, BD. | Department of Physiotherapy and Rehabilitation, JUST, Medical center room no-301 Jashore-7408, Bangladesh.<br>Not Applicable<br>N/A | 01778315139<br><br> |

**Details of Ethics Committee**

| Name of Committee | Approval Status | Date of Approval | Is Independent Ethics Committee? |
|---|---|---|---|
| Institutional Review Board, Department of Physiotherapy and Rehabilitation, JUST | Approved | 03/04/2024 | No |

**Regulatory Clearance Status from DCGI**

| Status | Date |
|---|---|
| Not Applicable | No Date Specified |

**Health Condition / Problems Studied**

| Health Type | Condition |
|---|---|
| Patients | Plantar fascial fibromatosis |

**Intervention / Comparator Agent**

| Type | Name | Details |
|---|---|---|
| Intervention | Structural Diagnosis and Management (SDM) Approach | Calf muscle release in prone position, in the direction of (superior to inferior, inferior to superior, lateral to medial, medial to lateral). That will complete in the three stage (stage-1: apply gental pressure; stage-2: apply firm pressure; stage-3: deep pressure). This pressure depends on the patient’s tolerance with the dosages of 6 repetition of each direction each duration 5 minutes with 2sets. Calf muscle release in rolling technique in |

| | | |
|---|---|---|
| | | supine position with the dosages of 10 rolls of the affected foot with 2 sets. Gastrocnemii and soleus muscles stretch (following the grade-1, 2, and 3). That will complete in three stage (stage-1:dorsiflexion in first one-third; stage-2:dorsiflexion up to middle one-third range; stage-3: dorsiflexion to the full range)with the dosages of 5 to 7 repetitions with 60 seconds hold with 2 sets. Myo-neural stretching in supine lying with 60 seconds hold with 2 set. 3 sessions per week, Total 12 sessions, 4 weeks. |
| Comparator Agent | Myofascial Release (MFR) | Deep stripping on the plantar fascia surface toward the calcaneus with the dosages of (5-7) repetitions with (15-30) seconds hold ewith 1 set. Deep stripping to the triceps surae (gastrocnemius, soleus and plantaris) part of posterosuperficial compartment of the calf with the dosage of (5-7) repetitions with (15-30) seconds hold with 1 set. Deep stripping to lateral compartment of the calf with the dosages of (5-7) repetitions with (15-30) seconds hold with 1 set. 3 sessions per week, Total 12 sessions, 4 weeks. |

**Inclusion Criteria**

| Inclusion Criteria | |
|---|---|
| **Age From** | 30.00 Year(s) |
| **Age To** | 70.00 Year(s) |
| **Gender** | Both |
| **Details** | 1. Planter heel pain in diabetic patients<br/> 2. Patients who willing to participate<br/> 3. Age between (30 to 70 years)<br/> 4. Both gender (male and female)<br/> 5. Patients with a diagnosis of unilateral or bilateral plantar heel pain, plantar fasciitis, and heel spur.<br/> 6. Planter heel pain is associated with plantar fasciitis and calcaneal spur, during the morning<br/> it’s very painful when taking place the foot is on the floor among diabetic patients, and pain<br/> during a long time of standing, after walking and walking on the floor with barefoot.<br/> 7. Plantar heel pain with calf muscle tightness and stress on perifascial structure.<br/> 8. Any traumatic history (not fracture of the foot)<br/> 9. Limited ankle dorsiflexion and activity limitation. |

**Exclusion Criteria**

| Exclusion Criteria | |
|---|---|
| **Details** | 1. Patients with active foot ulceration or infection, severe peripheral vascular disease.<br>2. Patient with contraindications to manual therapy or exercise interventions.<br>3. Diagnosis of severe osteoporosis, infection, tumor, gangrene, |

| | | |
|---|---|---|
| | | cyanotic symptom.<br>4. Patient who have any surgery of the foot. |

| | |
|---|---|
| **Method of Generating Random Sequence** | Computer generated randomization |
| **Method of Concealment** | Sequentially numbered, sealed, opaque envelopes |
| **Blinding/Masking** | Participant and Outcome Assessor Blinded |
| **Primary Outcome** | **Outcome**: As primary outcome, Pain, Disability, Activity limitation.<br>**Timepoints**: 12 Sessions,4 Weeks |
| **Secondary Outcome** | **Outcome**: As secondary outcome, Muscle strength & Range of Motion (ROM)<br>**Timepoints**: 12 Sessions,4 Weeks |
| **Target Sample Size** | **Total Sample Size=**70<br>**Sample Size from India=**0<br>**Final Enrollment numbers achieved (Total)=**Applicable only for Completed/Terminated trials<br>**Final Enrollment numbers achieved (India)=**Applicable only for Completed/Terminated trials |
| **Phase of Trial** | N/A |
| **Date of First Enrollment (India)** | 15/11/2024 |
| **Date of First Enrollment (Global)** | 15/11/2024 |
| **Estimated Duration of Trial** | **Years=**0<br>**Months=**11<br>**Days=**0 |
| **Recruitment Status of Trial (Global)** | Open to Recruitment |
| **Recruitment Status of Trial (India)** | Not Applicable |
| **Publication Details** | N/A |
| **Brief Summary** | Planter heel pain is the most common musculoskeletal condition of the lower extremities characterized by plantar fascia pain, perfascial structure pain, in the morning placing the first step on the floor then feel pain most in the foot, during prolonged time standing or walking feel pain, it can be caused for associated risk factors such as (type-1 or type-2 diabetes, obesity, most time stay in weight-bearing position which is more tend to occur heel pain, calcaneal spur). Both the SDM approach and MFR are effective for plantar heel pain but in the previous study where the results showed that the SDM approach is effective for improving heel pain compared with MFR. This study protocol will determine the comparison of the Structural Diagnosis and Management (SDM) Approach and MyoFascial Release (MFR) for plantar heel pain in diabetic patients. |
