## Supplementary material for "Comparison of Structural Diagnosis and Management (SDM) Approach versus Myofascial Release (MFR) for Plantar Heel Pain in People with Diabetes Mellitus: A Study Protocol for a Multicentre Randomised Clinical Trial": EthicsApproval_PTR JUST_2025

### Institutional Review Board (IRB)
**Department of Physiotherapy & Rehabilitation**
**Jashore University of Science & Technology**
**mail:** **website:** https://ptr.just.edu.bd/


PTR-JUST/IRB/2025/03/192411 April 03, 2025

To
Paroshmoni Biswas Mim
Student ID: 192411
Department of Physiotherapy and Rehabilitation
Jashore University of Science & Technology (JUST)
Jashore – 7408, Bangladesh

Approval of documents for the study titled **"Comparison of Structural Diagnosis and Management (SDM) Approach versus Myofascial Release (MFR) for Plantar Heel Pain in People with Diabetes Mellitus: A Multicenter Randomized Clinical Trial"** by the Institutional Review Board (IRB).

Dear Paroshmoni Biswas Mim,
Congratulations.
The Institutional Review Board (IRB) of the Department of Physiotherapy & Rehabilitation at Jashore University of Science & Technology has reviewed and discussed your application to conduct the research entitled **"Comparison of Structural Diagnosis and Management (SDM) Approach versus Myofascial Release (MFR) for Plantar Heel Pain in People with Diabetes Mellitus: A Multicenter Randomized Clinical Trial"** on the first IRB meeting held on March 30, 2025. We have reviewed the latest version of the Ethical Statement Checklist, Participant Information Sheet (PIS), Consent Form, and Questionnaire. **The IRB members are satisfied with your research proposal presentation held on March 30, 2025.**

**We approved the research to be conducted in its presented form with the following clauses-**

I. We confirm that neither you nor your study team members participated in the deliberations of the ethics committee & did not vote on the proposal for this study.
II. The institutional ethics committee expects to be informed about the progress of the study, any serious adverse effects (SAE) occurring in the course of the study, any changes in the protocol and participant's information/ informed consent and asks to be provided a copy of the final record.
III. Please submit the published article of the study as per EC standard operating protocols.
IV. The EC is organized and operates according to the requirements of the declaration of the Helsinki and ICH-GCP, local regulatory requirements, and guidelines.
V. The study is partially funded by Jashore University of Science and Technology, Jashore-7408, Bangladesh.

Yours sincerely,

**Dr. Md. Shahadat Hossain**
Assistant Professor (Adjunct), Dept. of Physiotherapy and Rehabilitation
Member Secretary, Institutional Review Board (IRB)
Jashore University of Science and Technology,
Jashore-7408, Bangladesh.
